# Immune responses and disease biomarker long-term changes following COVID-19 mRNA vaccination in a cohort of rheumatic disease patients

**DOI:** 10.1101/2023.03.22.23287597

**Authors:** Zesheng An, Xian Zhou, Yanfeng Li, Jane Jaquith, Kathleen McCarthy-Fruin, Jennifer Sletten, Kenneth J. Warrington, Cornelia Weyand, Cynthia S. Crowson, Saranya Chumsri, Keith L. Knutson, Gabriel Figueroa-Parra, Alain Sanchez-Rodriguez, Uma Thanarajasingam, Alí Duarte-García, Hu Zeng

**Affiliations:** Division of Rheumatology, Department of Medicine, Mayo Clinic Rochester, MN 55905, USA; Department of Urology, Tianjin Institute of Urology, The Second Hospital of Tianjin Medical University, Tianjin, P. R. China, 300211; Department of Cancer Biology, Mayo Clinic, Jacksonville, FL 32224, USA; Department of Quantitative Health Sciences, Mayo Clinic Rochester, MN 55905, USA; Department of Immunology, Mayo Clinic, Jacksonville, FL 32224, USA; Department of Immunology, Mayo Clinic Rochester, MN 55905, USA

**Keywords:** COVID-19 mRNA vaccine, systemic lupus erythematosus, Sjögren’s syndrome, Psoriatic arthritis

## Abstract

**Objective:** To evaluate seroreactivity and disease biomarkers after 2 or 3 doses of COVID-19 mRNA vaccines in a cohort of patients with rheumatic diseases.

**Methods:** We collected biological samples longitudinally before and after 2-3 doses of COVID-19 mRNA vaccines from a cohort of patients with systemic lupus erythematosus (SLE), psoriatic arthritis, Sjogren’s syndrome, ankylosing spondylitis, and inflammatory myositis. Anti-SARS-CoV-2 spike IgG and IgA and anti-dsDNA concentration were measured by ELISA. A surrogate neutralization assay was utilized to measure antibody neutralization ability. Lupus disease activity was measured by Systemic Lupus Erythematosus Disease Activity Index (SLEDAI). Expression of type I interferon signature was measured by real-time PCR. The frequency of extrafollicular double negative 2 (DN2) B cells was measured by flow cytometry.

**Results:** Most of the patients generated high SARS-CoV-2 spike-specific neutralizing antibodies comparable to those in healthy controls after 2 doses of mRNA vaccines. The antibody level declined over time but recovered after the third dose of the vaccine. Rituximab treatment substantially reduced antibody level and neutralization ability. Among SLE patients, no consistent increase in SLEDAI scores was observed post-vaccination. The changes in anti-dsDNA antibody concentration and expression of type I IFN signature genes were highly variable but did not show consistent or significant increases. Frequency of DN2 B cells remained largely stable.

**Conclusion:** Rheumatic disease patients without rituximab treatment have robust antibody responses toward COVID-19 mRNA vaccination. Disease activity and disease-associated biomarkers remain largely stable over 3 doses of vaccines, suggesting that COVID-19 mRNA vaccines may not exacerbate rheumatic diseases.

**KEY MESSAGES:**

- Patients with rheumatic diseases mount robust humoral immunity towards 3 doses of COVID-19 mRNA vaccines.
- Disease activity and biomarkers remain stable following 3 doses of COVID-19 mRNA vaccines.

## INTRODUCTION

The coronavirus disease 2019 (COVID-19) messenger RNA (mRNA) vaccines have achieved remarkable success in fighting the COVID-19 pandemic caused by SARS-CoV-2. Because patients receiving immunosuppressive medicines were excluded in initial clinical trials and were less represented in many subsequent longitudinal studies, our understanding of the longitudinal immune responses towards COVID-19 mRNA vaccines in these patients remains incompletely understood. Furthermore, although several studies have indicated that COVID-19 mRNA vaccines usually do not exacerbate existing systemic autoimmunity, the current data are mostly observational, often over a short period (e.g., a few days after 2^nd^ dose of vaccine) and may not account for the potential latent time for disease flare (1-5). Because changes in many serological, cellular, and molecular markers can precede disease onset, we reason that these biomarkers may provide a better indication of whether COVID-19 mRNA vaccines have any impact on disease activity. One consideration is that overactivation of type I interferon (IFN) has been associated with multiple rheumatic diseases, including systemic lupus erythematosus (SLE) and Sjögren’s syndrome. mRNA vaccines are known to provoke type I IFN signaling (6). Thus, it is plausible that repeated mRNA vaccination might lead to disease exacerbation partly due to type I IFN activation. To address this question, we evaluated disease activity, immune responses, cellular compositions, autoantibody, and type I IFN signatures in samples from a cohort of rheumatic disease patients longitudinally collected over 2-3 doses of COVID-19 mRNA vaccines. Our data show that repeated COVID-19 mRNA vaccination does not significantly increase disease activity scores, autoantibody level, expression of type I IFN signature genes, and immune subsets associated with systemic autoimmunity. Thus, these results suggest that repeated COVID-19 mRNA vaccination may not lead to increase disease activity in rheumatic disease patients.

## PATIENTS AND METHODS

### Study population and sample collection/storage

Patients were recruited from the Division of Rheumatology at Mayo Clinic Rochester in 2021. The study has been approved by Institutional Review Board of Mayo Clinic Rochester (IRB 21-000501). Written informed consent was obtained from all participating patients. Patients’ demographics, clinical information and medication exposures can be found in **Supplementary Table 1 and Table 2**. All 28 patients received at least two doses of COVID-19 mRNA vaccines (either Pfizer or Moderna mRNA vaccines). Among them, 13 received three doses of COVID-19 mRNA vaccination. Healthy controls’ plasma was collected by one month and six months after their 2^nd^ dose vaccination at Mayo Clinic Rochester or Florida. Unvaccinated healthy control plasma was collected before the COVID-19 pandemic. Patients’ whole blood was collected at Visit 1 (before the 1^st^ dose), Visit 2 (1 month after the 2^nd^ dose), Visit 3 (4 months after the 2^nd^ dose), and Visit 4 (1 month after the 3^rd^ dose of COVID-19 mRNA vaccination). Whole blood samples were preserved in DNA/RNA Shield (Zymo Research) and stored at -80°C. Peripheral blood mononuclear cells (PBMC) were isolated using Ficoll density gradient centrifugation and were resuspended in freezing medium. Subsequently, PBMC were cryopreserved in liquid nitrogen until use.

### Enzyme-linked immunosorbent assay (ELISA) for recombinant SARS–CoV-2 spike protein

Ninety-six-well plates were coated with 1 μg/mL SARS-CoV-2 (2019-nCoV) Spike S1-His Recombinant Protein (Sino Biological) in PBS and incubated overnight at 4°C. Human plasma samples were heat-inactivated at 56°C for 1 hour. Plates were blocked with 3% Blotting-Grade Blocker (BIO-RAD) in 0.05% PBST for 1 hour at 37°C. Plasma samples were serially diluted in three-fold with 1% Blotting-Grade Blocker (BIO-RAD) in 0.05% PBS-T (dilution buffer), starting at 1:450 to 1:109350 dilution. SARS-CoV-2 Spike Protein (CR3022) Human IgG1 mAb (Cell Signaling) was included on each plate as Internal control to convert OD values into antibody concentrations. Diluted plasma samples were added to each well and incubated for 2 hours at room temperature using a plate mixer. Samples were run in duplicate. In each plate, convalescent plasma diluted samples were included as positive controls; unvaccinated healthy control diluted plasma samples were included as the negative control. Wells only added with dilution buffer were set in each plate as background readout. Detection relied on an enzyme-labeled secondary antibody, 1:10,000 diluted Goat Anti-Human IgG ads-HRP (Southern Biotech), or 1:1000 diluted Goat Anti-Human IgA ads-HRP (Southern Biotech) was added to each well for 1 hour at room temperature using a plate mixer. After incubation, TMB One Component HRP Microwell Substrate (Surmodics) was added to each well, and the reaction was terminated by 2M H_2_SO_4_. The absorbances were detected using a microplate reader at an optical density (OD) of 450 nm and 620 nm. The IgG original concentration and IgA half maximum dilution of the plasma samples were calculated and generated by GraphPad Prism 9.

### SARS-CoV-2 Surrogate Neutralization Assay

To detect SARS-CoV-2 total neutralizing antibodies and to identify individuals with an adaptive immune response to SARS-CoV-2, we used cPass SARS-CoV-2 Neutralization Antibody Detection Kit (GenScript) as a surrogate neutralization assay, which measures the ability of plasma to block the interaction between the ACE2 receptor protein and the receptor binding domain (RBD) of the viral spike protein. Convalescent plasma, healthy control plasma samples, and all 13 patients who have Visit 4 timepoint plasma samples were tested according to the manufacturer’s protocol. Human plasma samples were heat-inactivated at 56°C for 1 hour. Test plasma samples, positive and negative controls were diluted by sample dilution buffer with a ratio of 1:10. All plasma samples and controls were tested in duplicate. After reading the absorbance in a microtiter plate reader at 450 nm, the human plasma samples can be separated as a positive result and a negative result. Following the selection, the human plasma samples with a positive result were serially diluted in three-fold with sample dilution buffer, starting at 1:20 to 1:4860 dilution. The diluted plasma samples were tested with the same kit to obtain the titer measurement readout. The half-maximum dilution of the plasma samples with a positive result was calculated and generated by GraphPad Prism 9.

### Anti-double stranded deoxyribonucleic acid (dsDNA) ELISA assay

To measure the specific IgG autoantibodies against the double stranded deoxyribonucleic acid (dsDNA) in human plasma samples, we used QUANTA Lite dsDNA SC ELISA (Inova Diagnostics) to test patients’ samples according to the manufacturer’s protocols. Human plasma samples were diluted by the sample dilution buffer with a ratio of 1:100, as recommended. The calibrator samples, positive, negative, and single-stranded controls, were involved in each plate. Read the optical density (OD) of each well at 450nm on a microplate reader. The original concentration of the plasma samples was calculated by ratio.

### Whole blood RNA isolation and Real-Time PCR

Whole blood RNA was isolated using Quick-RNA Whole Blood (Zymo Research) according to the manufacturer’s instructions and protocols. RNA concentration was measured by spectrophotometry, and the sample’s purity was evaluated according to 260/280 nm ratio. And cDNA was generated from RNA through PrimeScript RT Master Mix (TaKaRa). Then, cDNA was amplified by Real-Time PCR (Applied Biosystems) using Applied ABI PowerUp™ SYBR™ Green Master Mix (Life Technologies) and specific primers for the interest genes (*MX1, IFIT1, IFI44*). All reactions were run in duplicate under the same thermal cycling conditions as follows: 95°C for 10 min (polymerase activation) followed by 40 cycles at 95°C for 30s, 52-60°C for 30s, and 72°C for 30s. The results were quantified using the comparative (2-ΔΔCt) method. The Ct value of the target gene was normalized to the Ct value of the housekeeping gene, β-actin. Each patient’s data were reported as fold increase of the target gene mRNA level compared to his/her Visit 1 sample. The primer sequences are listed in **Supplementary Table 3**.

### Flow cytometry

Cryopreserved PBMCs were thawed and stained with antibodies or viability dyes listed in **Supplementary Table 4**. Samples were acquired on a 4-laser Attune NxT flow cytometer (ThermoFisher Scientific). Up to 1.5 × 10^6^ PBMCs were acquired for each sample. All samples were acquired with consistent settings on the same day. Data were analyzed with FlowJo 10.8.1 (Becton Dickinson).

### Lupus disease activity estimate

SLE disease activity was estimated retrospectively through the SLE disease activity index (SLEDAI) at the time of the vaccination by two rheumatologists (7).

### Statistical analysis

The statistical analysis was performed using the GraphPad Prism 9 software (GraphPad). Continuous variables were compared using the unpaired and paired Student T-test. P-values < 0.05 were considered statistically significant.

## RESULTS

### Robust humoral immune response to COVID-19 mRNA vaccine in rheumatic patients

Although recent literature indicates that some patients with rheumatic diseases had modestly reduced humoral immune response following two doses of COVID-19 mRNA vaccination, there was a considerable variation (2, 8). We first sought to determine if our cohort of rheumatic patients had good immune responses toward the COVID-19 mRNA vaccines (Moderna/Pfizer). To this end, we longitudinally measured human IgG and IgA antibodies against the SARS-CoV-2 spike protein S1 subunit at different time points before and after vaccination. Throughout our study, Visit 1 (V1) represents “before the 1^st^ dose of vaccination”; Visit 2 (V2) represents “1 month after the 2^nd^ dose vaccination”; Visit 3 (V3) represents “4 months after the 2^nd^ dose vaccination”; Visit 4 (V4) represents “1 month after the 3^rd^ dose vaccination”. Sex and age-matched healthy control samples collected before the COVID-19 pandemic, one month after the 2^nd^ vaccination or six months after the 2^nd^ vaccination, and convalescent samples after recovering from COVID-19 were included as controls.

We separated our cohort of rheumatic disease patients into three groups (**Figure 1**). Orange lines denote patients diagnosed with SLE, Sjogren’s, and idiopathic inflammatory myopathies, which are considered systemic autoimmune diseases. Green lines denote patients diagnosed with psoriatic arthritis and ankylosing spondylitis, which are considered inflammatory arthritides. Blue lines indicate two rituximab-treated patients, one diagnosed with SLE and the other with Sjogren’s. Rituximab-treated patients were separated because rituximab is known to have significantly negative effects on the humoral immune response to SARS-CoV-2 mRNA vaccines (2, 9-11).

**Figure 1.**
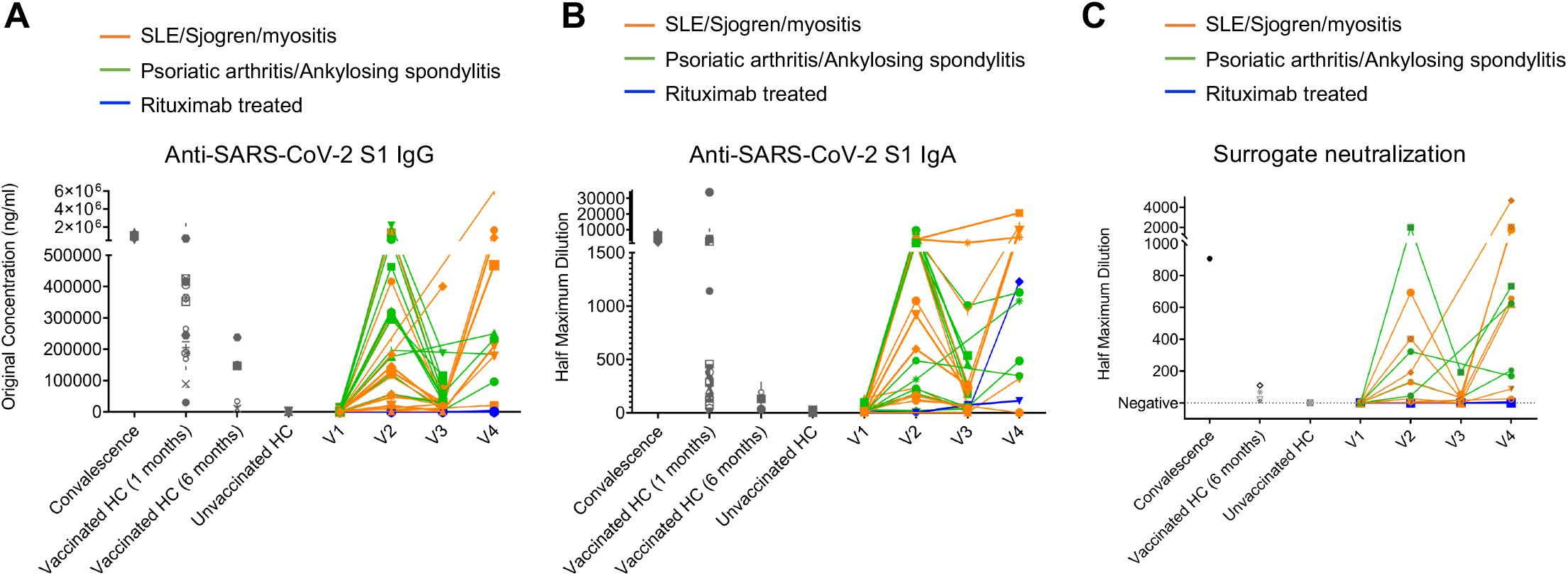
Humoral response to COVID-19 mRNA vaccine in rheumatic disease patients. Throughout, orange lines represent patients confirmed with SLE, Sjogren, and Myositis. Green lines represent patients confirmed with Psoriatic arthritis and Ankylosing spondylitis. Blue lines represent two patients treated with rituximab confirmed with SLE and Sjogren, respectively. **A**, Concentration of human IgG against the SARS-CoV-2 spike S1 protein was obtained by ELISA. Orange subgroup (n = 14). Green subgroup (n = 12). Blue subgroup (n = 2). Healthy controls (HC) post-vaccinated 1-month (n = 20). HC post-vaccinated 6-month (n = 5). Unvaccinated healthy controls (HC) (n = 7). **B**, Quantification of human IgA titers against the SARS-CoV-2 spike S1 protein was obtained by ELISA. **C**, The ability of serum antibodies to block SARS-CoV-2 binding to human angiotensin-converting enzyme-2 (hACE2) receptor measured by surrogate neutralization.

Our ELISA results showed that anti-SARS-CoV-2 spike S1 IgG levels increased from barely detectable pre-vaccination to an equivalent level as those in vaccinated healthy controls (HC) at V2 (1 month after 2^nd^ dose). Then they declined at V3 (4 months after 2^nd^ dose). At V4 (after the 3^rd^ dose), the IgG levels recovered to a similar level compared to those at V2 (**Figure. 1A and Supplementary Figure 1A, 1B**). The exceptions are those receiving rituximab whose anti-SARS-CoV-2 spike S1 IgG levels remained undetectable throughout all visits, consistent with previous reports (9, 11-13) (**Figure. 1A**). IgA provides vital immune protection, especially in the mucosal sites. The dynamics of anti-SARS-CoV-2 spike S1 IgA titers broadly resembled those of anti-SARS-CoV-2 spike S1 IgG, except that the two rituximab-treated patients unexpectedly mounted measurable levels of S1 specific IgA at V4 (**Figure 1B and Supplementary Figure 1C**). The reduction of IgG and IgA levels at V3 compared to V2, and the recoveries at V4 compared to V3, were mostly statistically significant (**Supplementary Figure 1A, 1B**).

Furthermore, we evaluated the serum neutralizing ability using a surrogate angiotensin converting-enzyme 2 (ACE2) binding assay, which measures the ability of antibodies in plasma to block the interaction between the human angiotensin-converting enzyme-2 (hACE2) receptor protein and the receptor binding domain (RBD) of the viral spike protein. The data revealed similar dynamics as in the serum IgG and IgA titer results. The patient V2 samples showed a robust ability to block ACE2 binding, which declined at V3 and recovered at V4. We did not detect any neutralizing ability in the samples from the two rituximab-treated patients, suggesting that the measurable anti-SARS-CoV-2 S1 IgA in these two patients did not have the neutralizing capability (**Figure 1C**). Statistical analyses showed that patients with rheumatic diseases (excluding the rituximab-treated patients) generated anti-SARS-CoV-2 spike S1 IgG and IgA one month after 2^nd^ dose at a level comparable to healthy control groups (**Supplementary Figure 1B, 1D**). While the antibodies waned at V3, they recovered to a similar (IgG) or slightly higher level (IgA) as in V2 after the 3^rd^ dose (**Supplementary Figure 1A, 1C**). There was no significant difference between the systemic autoimmune disease group and the inflammatory arthritis group at each visit (**Supplementary Figure 2**). Together, these data showed that patients with rheumatic diseases had a robust humoral immune response towards COVID-19 mRNA vaccines after two doses, followed by a gradual decline and a strong recovery of anti-SARS-CoV-2 S1 IgG and IgA titers after a 3^rd^ dose. Patients treated with rituximab had negligible SARS-CoV-2 specific IgG titers in all visits and some measurable level of IgA after the 3^rd^ dose of vaccination. However, these IgA might not confer neutralization capability.

### Repeated SARS-CoV-2 vaccinations do not have a significant impact on multiple disease biomarkers

One of the outstanding questions is the safety of COVID-19 mRNA vaccines in rheumatic disease patients. Autoimmune responses can be triggered by immune challenges, including vaccination, evidenced by rare myocarditis following COVID-19 mRNA vaccination in young adults (14). To assess the clinical symptom severity, we first assessed the progress of clinical disease activity, measured by Systemic Erythematosus Disease Activity Index (SLEDAI), among the patients with lupus in our cohort. We found that 3 out of 12 patients showed modestly increased SLEDAI scores at the last visit compared to their first visit. The SLEDAI scores for the other patients remained either stable or modestly reduced (**Figure. 2**). Thus, we did not observe a widespread increase in lupus disease activity after COVID-19 mRNA vaccination.

**Figure 2.**
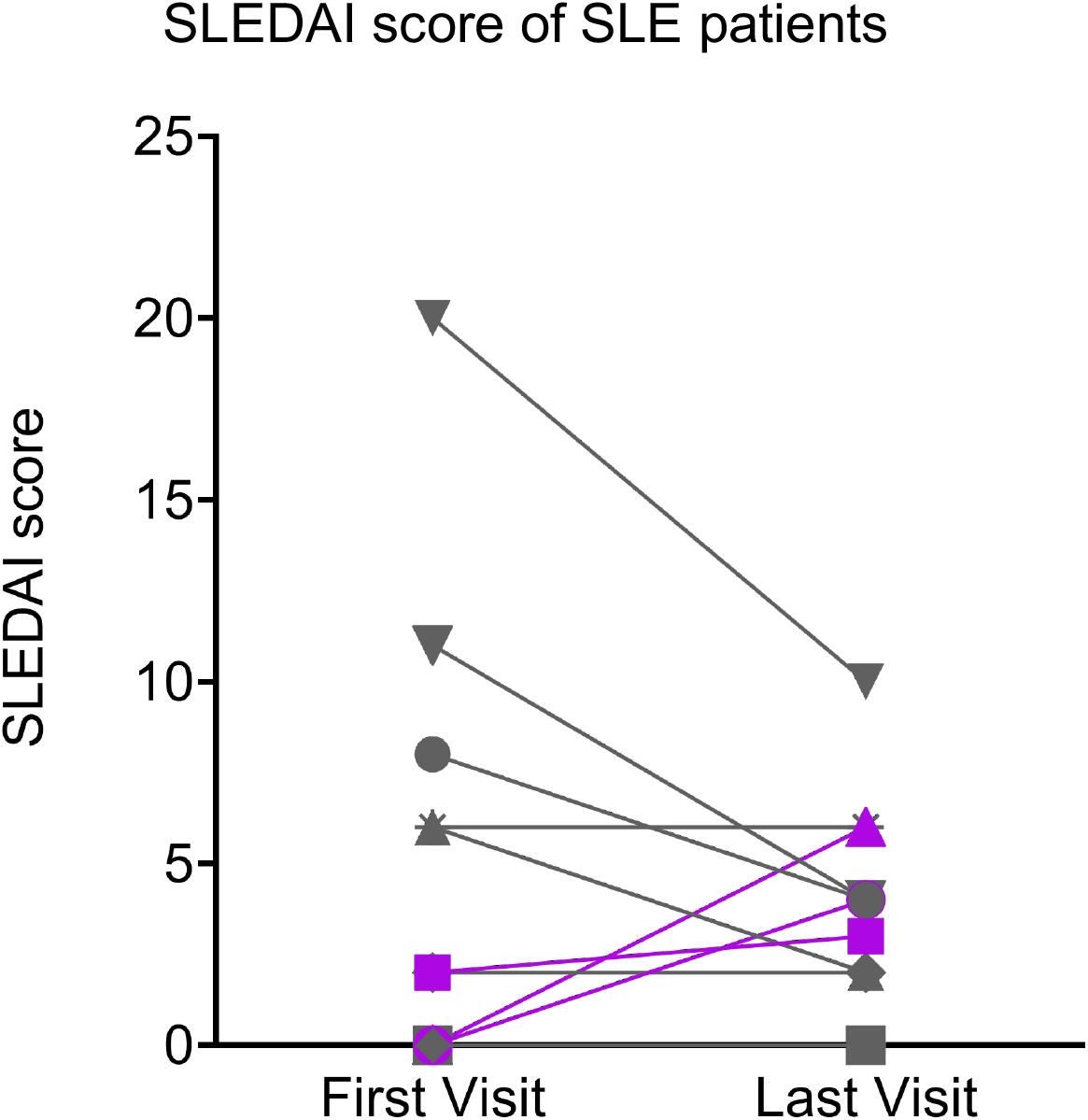
The Systemic Erythematosus Disease Activity Index (SLEDAI) of SLE patients (n=12) was obtained at their first visits and last visits. Gray lines represent the SLE patients (n=9) with decreasing or stable SLEDAI scores, and purple lines represent the SLE patients (n=3) with increasing SLEDAI scores at the last visits compare to their first visits.

Because many molecular and cellular phenotypes can long precede autoimmune disease onset or flare, we decided to evaluate several key biomarkers associated with systemic rheumatic diseases. These included anti-dsDNA concentration, expression of type I IFN signature genes, and the frequency of IgD and CD27 ‘double-negative’ (DN) B cells lacking CD21 expression but expressing CD11c (DN2 cells), which expanded in patients with active SLE and correlated with increased morbidity in COVID-19 patients (15, 16). Plasma anti-dsDNA concentration was measured with ELISA. Samples from vaccinated healthy donors were included as controls. As expected, SLE patients had relatively high levels of anti-dsDNA autoantibodies, while vaccinated healthy controls had the lowest level of anti-dsDNA autoantibodies (**Figure 3A**). When we compared anti-dsDNA levels between V1 and each subsequent visit, we did not observe a consistent increase of anti-dsDNA in any comparison (**Figure 3B**). Thus, 2 or 3 doses of COVID-19 mRNA vaccines do not substantially affect anti-dsDNA antibody levels in SLE patients.

**Figure 3.**
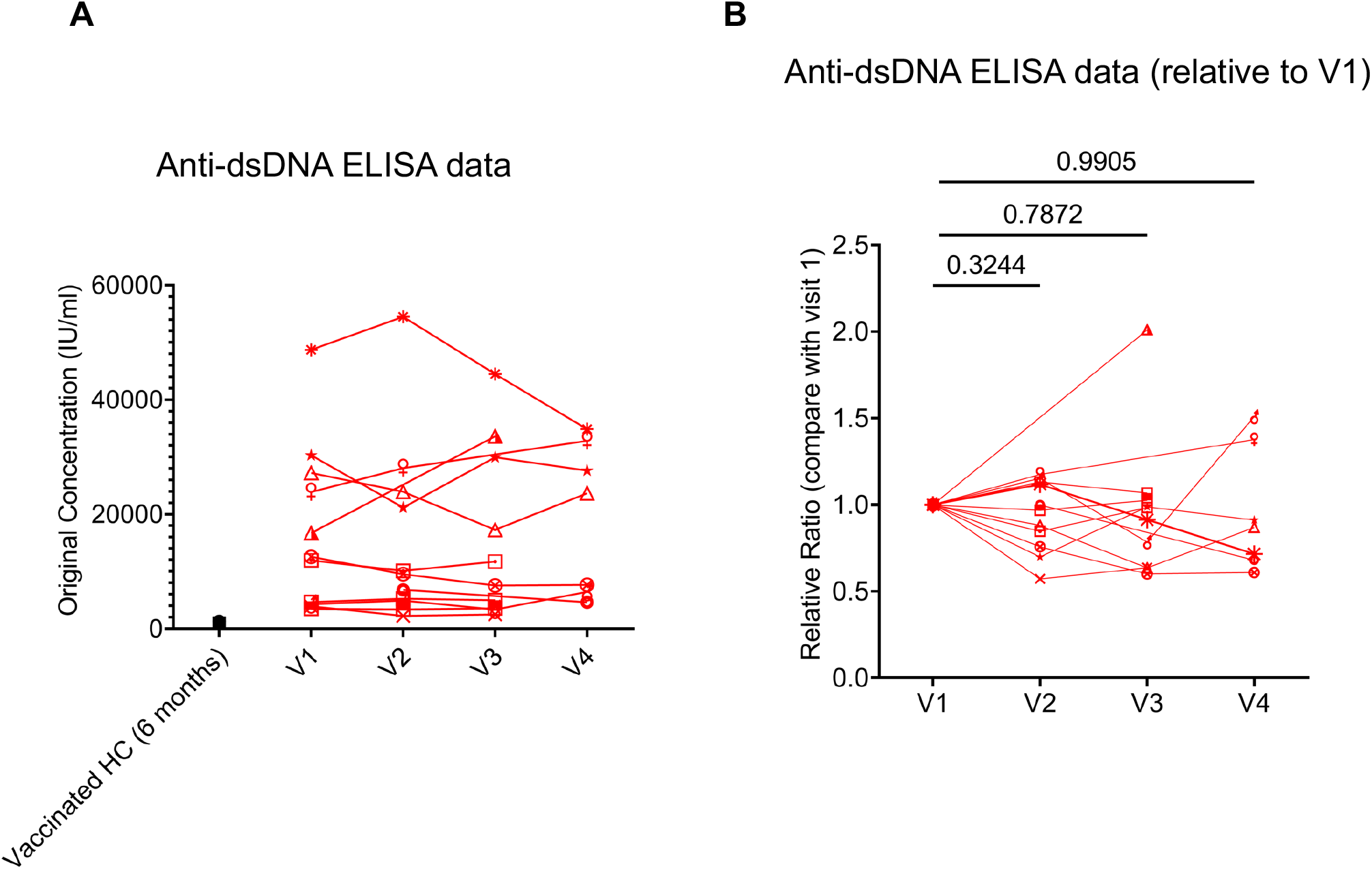
Human plasma anti-double stranded deoxyribonucleic acid (anti-dsDNA) level. **A**, Concentration of anti-dsDNA in human plasma samples. SLE disease group (Red, n = 11). Non-SLE disease group (Grey, n = 17). Healthy controls post-vaccinated six months (n = 5). Unvaccinated healthy control (n = 2). **B**, Plasma anti-dsDNA antibody levels relative to those in V1. *P* values were calculated with paired parametric t-test.

Previous literature has demonstrated that COVID-19 mRNA vaccines can stimulate serum type I IFN production, which is critical for optimal cellular immune response to the vaccine (6). On the other hand, there is evidence implicated that high mRNA transcripts of genes regulated by type I IFN, also known as type I IFN signatures, are correlated with the disease activity and the pathogenesis of a number of systemic autoimmune diseases, including SLE, Sjogren’s syndrome, rheumatoid arthritis and myositis (17). We examined the mRNA expression of three type I IFN signature genes (*MX1, IFIT1*, and *IFI44*) (18) in whole blood samples by real-time PCR. We observed a trend of increased *IFI44* and *IFIT1* expression at V2 relative to V1. At V3 and V4, none of the three genes showed any trend of increased expression compared to V1. In fact, the expression of *IFI44* at V4 was significantly reduced compared to V1 (**Figure 4**). Therefore, 2 or 3 doses of COVID-19 mRNA vaccines do not consistently increase the expression of type I IFN signature genes.

**Figure 4.**
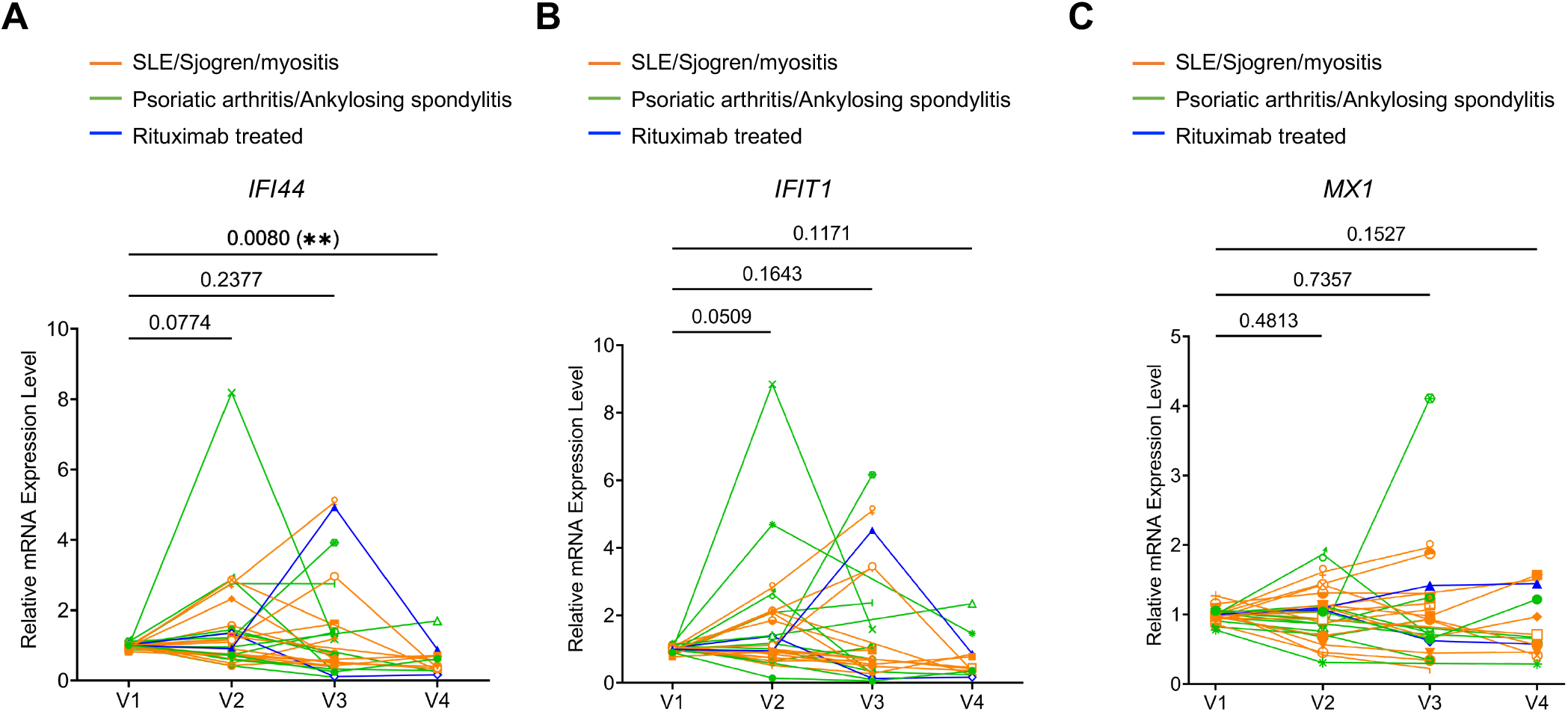
Expression of type I interferon (IFN) signature genes in rheumatic disease patients’ whole blood following SARS-CoV-2 mRNA vaccination. Orange subgroup (n = 14). Green subgroup (n = 12). Blue subgroup (n = 2). Relative mRNA expression of *IFI44* (**A**), *IFIT1* (**B**), and *MX1* (**C**) were measured by RT-qPCR. *P* values were derived from a paired parametric t-test to show comparisons between indicated groups.

Finally, we sought to evaluate the cellular changes following COVID-19 mRNA vaccination using flow cytometry (see **Supplementary Figure** 3 for gating strategy). For non-rituximab-treated patients, the frequency of CD19^+^ B cells showed a general trend of slight reduction relative to V1 (reached statistical significance only at V3). The two patients who were treated with rituximab had CD19^+^ frequencies below 0.02% at every visit, confirming the efficient B cell depletion (**Supplementary Figure 4A**). Within B cells, CD27^+^CD38^+^ plasmablasts remained stable throughout all visits (**Supplementary Figure 4B**). Extrafollicular DN2 B cells (CD19^+^IgD^−^ CD27^−^CD11c^+^CD21^−^) are associated with systemic autoimmunity. We found that DN2 B cell frequency did not show any significant alterations. The two rituximab-treated patients showed highly increased DN2 B cell frequency at V3 (**Figure 5**). However, due to the extremely low CD19^+^ frequency, the biological impact of such changes was likely limited. Moreover, examination of T cells showed that total CD3^+^ T cell frequency, but not CD4^+^ or CD8^+^ T cell frequencies, slightly increased over the course of vaccination (**Supplementary Figure 4C-E**). The specialized B cell-interacting follicular helper T cells (Tfh)-like cells (CD4^+^CD45RA^−^CXCR5^+^) remained stable (**Supplementary Figure 4F**). Thus, overall lymphocyte homeostasis, including autoimmunity-associated extrafollicular DN2 B cells, is not substantially altered by COVID-19 mRNA vaccination in rheumatic disease patients.

**Figure 5.**
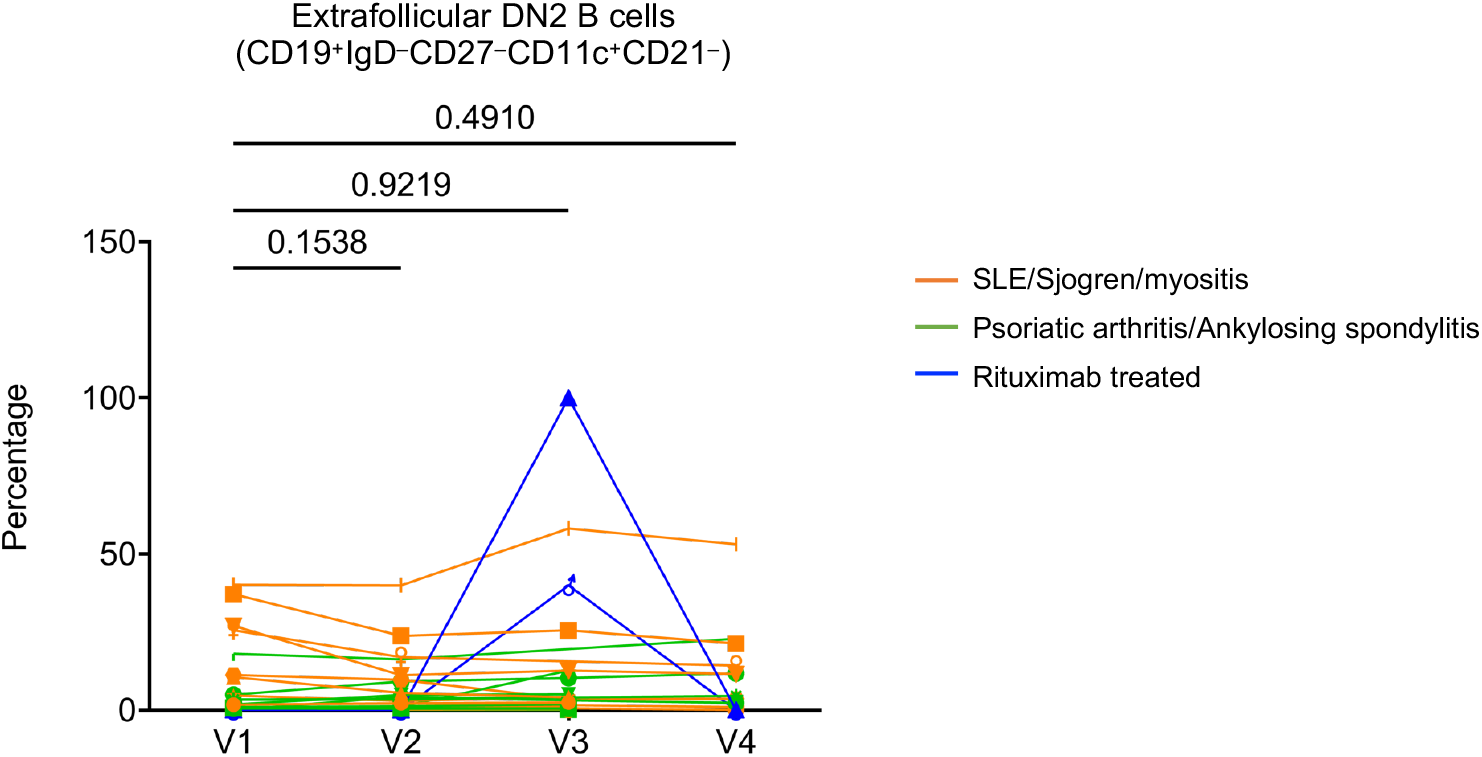
The frequencies of Extrafollicular DN2 B cells (CD19^+^IgD^−^CD27^−^CD11c^+^CD21^−^) following SARS-CoV-2 mRNA vaccination in patients’ PBMC. Orange subgroup (n = 14). Green subgroup (n = 12). Blue subgroup (n = 2). *P* values were generated by paired parametric t-test.

## DISCUSSION

Our study shows that following two doses of COVID-19 mRNA vaccines, most of the patients with rheumatic diseases elicit robust humoral immunity. The anti-SARS-CoV-2 antibody titers and neutralization ability declined over time after 2^nd^ dose of the vaccine but mostly recovered after a 3^rd^ dose of the COVID-19 mRNA vaccine. Previous studies have mostly focused on immune responses after two doses of COVID-19 vaccines, most of which reported a slightly reduced antibody response (1, 8). If the patients were treated with rituximab, their humoral response to COVID-19 mRNA vaccines was severely curtailed or even abolished (9, 11-13). Our data confirmed the detrimental effects of rituximab on the humoral response to COVID-19 mRNA vaccines. We did not observe a substantial reduction of anti-SARS-CoV-2 immunoglobulin in our patient cohort compared to healthy controls. Moreover, our data show that most patients can mount a vigorous antibody response after a 3^rd^ vaccination, which restores the antibody level to a similar or even higher level to the one observed one month after 2^nd^ dose. Thus, our results support that COVID-19 mRNA vaccines are highly immunogenic in patients with rheumatic diseases.

We also provide the first molecular and cellular evidence supporting the safety of long-term use of the COVID-19 mRNA vaccine in systemic rheumatic disease patients. While clinical observations support the safety of COVID-19 mRNA vaccines in rheumatic disease patients, anecdotal reports indicate a possible link between COVID-19 mRNA vaccines and disease flare (19). Therefore, we assessed the potential impacts of 2 or 3 doses of COVID-19 mRNA vaccines on disease activity and various disease activity biomarkers, including anti-dsDNA antibody level, expression of type I IFN signature genes, and cellular components. Reassuringly, the data showed that neither clinical disease activity (measured by SLEDAI in lupus patients) nor biomarkers indicate increased disease exacerbation following three doses of COVID-19 mRNA vaccines. Our findings corroborate the current clinical observations that COVID-19 mRNA vaccines are generally safe for rheumatic disease patients (4). Furthermore, at the time of preparing this manuscript, people in the US are getting updated bivalent COVID-19 mRNA vaccines as a 4^th^ dose. Our data suggest that such repeat mRNA vaccination might not increase the risk of disease exacerbation.

The major limitation of our study is the small cohort size, which precludes a definitive conclusion. Research with a larger cohort is warranted to verify our observations. Nevertheless, our study provides empirical and experimental evidence affirming the efficacy and safety of COVID-19 mRNA vaccines and supporting their wide usage in patients with systemic autoimmunity.

## Data Availability

All data produced in the present study are available upon reasonable request to the authors

## ACKNOWLEDGMENTS

The authors would like to thank the patients who participated in the study.

## AUTHOR CONTRIBUTIONS

**Study conception and design**. Zesheng An, Kenneth J. Warrington, Cornelia Weyand, Uma Thanarajasingam, Alí Duarte García, Hu Zeng.

**Acquisition of data**. Zesheng An, Xian Zhou, Yanfeng Li, Jane Jaquith, Kathleen McCarthy-Fruin, Jennifer Sletten, Saranya Chumsri, Keith L. Knutson, Gabriel Figueroa-Parra, Alain Sanchez-Rodriguez.

**Analysis and interpretation of data**. Zesheng An, Xian Zhou, Cynthia Crowson, Alí Duarte García, Hu Zeng

---

**Supplementary Figure 1.**
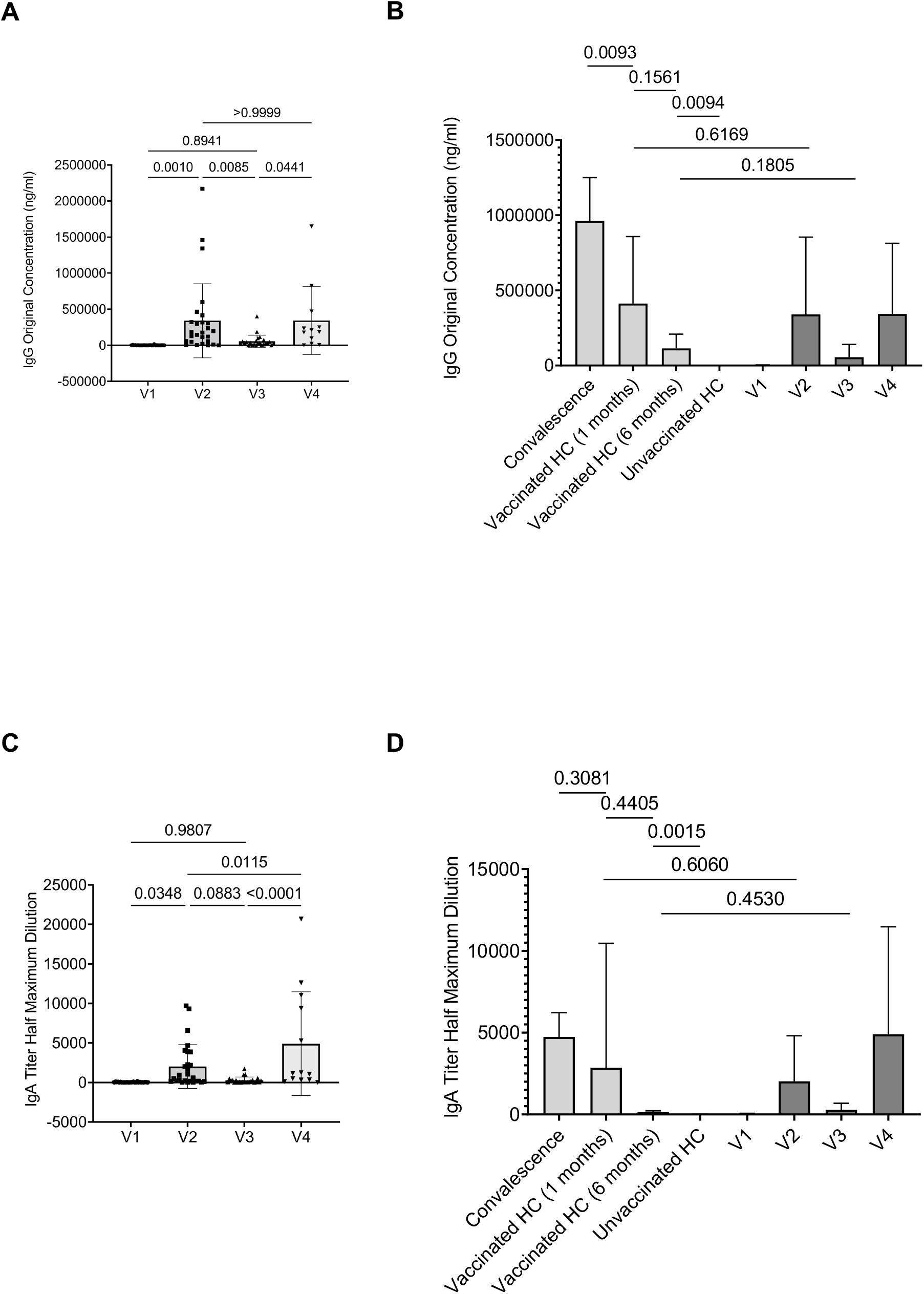
Statistical analyses of antibody titer data between visits. **A**, Comparisons of IgG concentration in rheumatic disease patients’ plasma samples between different visits. **B**, Concentration of human IgG against the SARS-CoV-2 spike S1 protein was obtained by ELISA. **C**, Comparisons of IgA titers half maximum dilution in rheumatic disease patients’ plasma samples between different visits. **D**, Quantification of human IgA against the SARS-CoV-2 spike S1 protein was obtained by ELISA. P values were derived from paired t-tests (A, C) and unpaired t-tests.

**Supplementary Figure 2.**
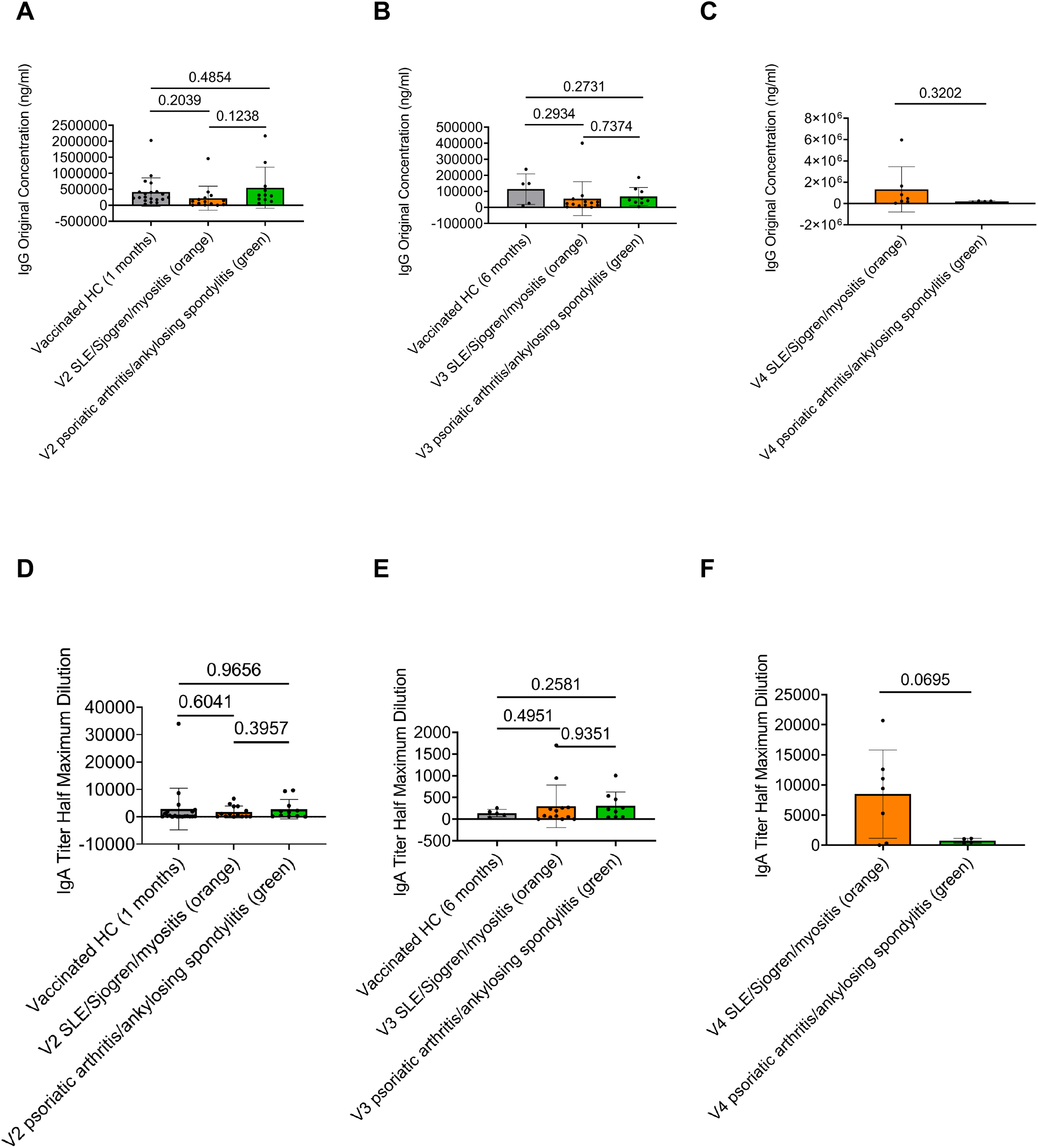
Statistical analyses of antibody titer between disease groups at different visits. **A-C**, Comparisons of anti-SARS-CoV-2 S1 protein IgG concentration between HC, SLE/Sjogren/Myostitis, and Psoriatic arthritis/Ankylosing spondylitis at V2 (**A**), V3 (**B**) and V4 (**C**). **D-F**, Comparisons of anti-SARS-CoV-2 S1 protein IgA titers between HC, SLE/Sjogren/Myostitis, and Psoriatic arthritis/Ankylosing spondylitis at V2 (**D**), V3 (**E**) and V4 (**F**). *P* values were calculated by unpaired parametric t-test.

**Supplementary Figure3.**
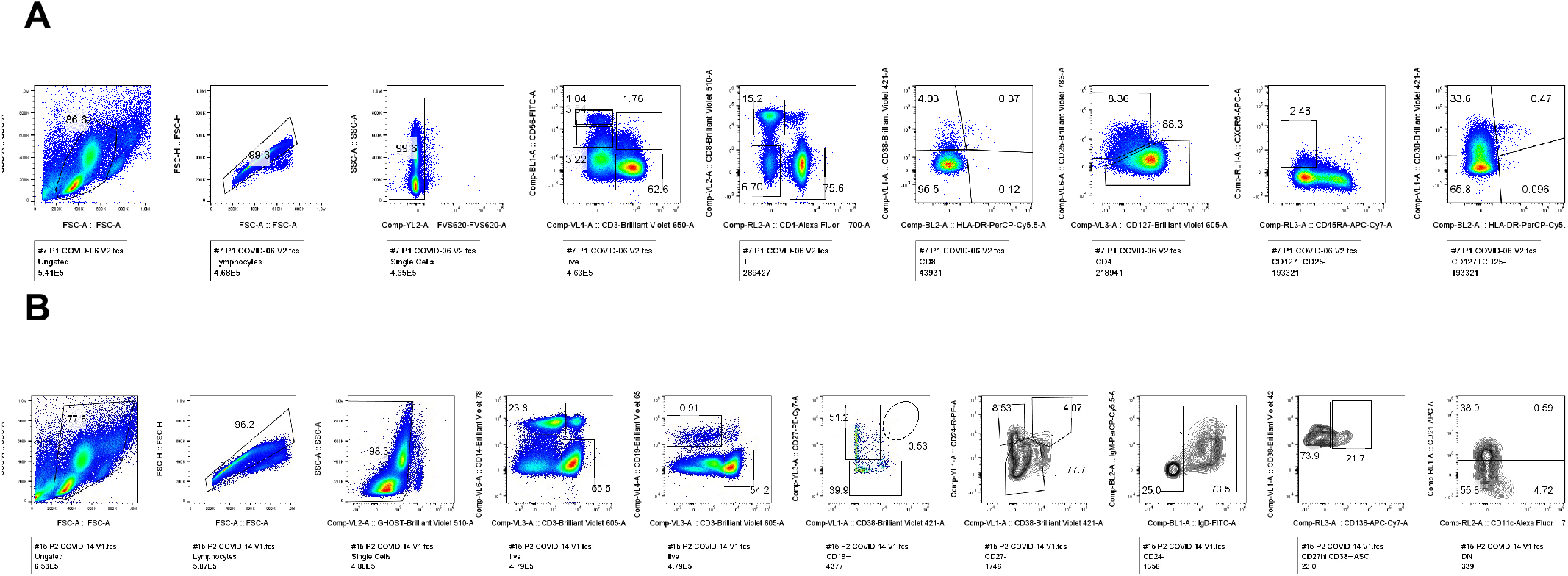
Representative flow cytometry plots of the gating strategies. **A**, Gating strategies for T cell subsets. **B**, Gating strategies for B cell subsets.

**Supplementary Figure4.**
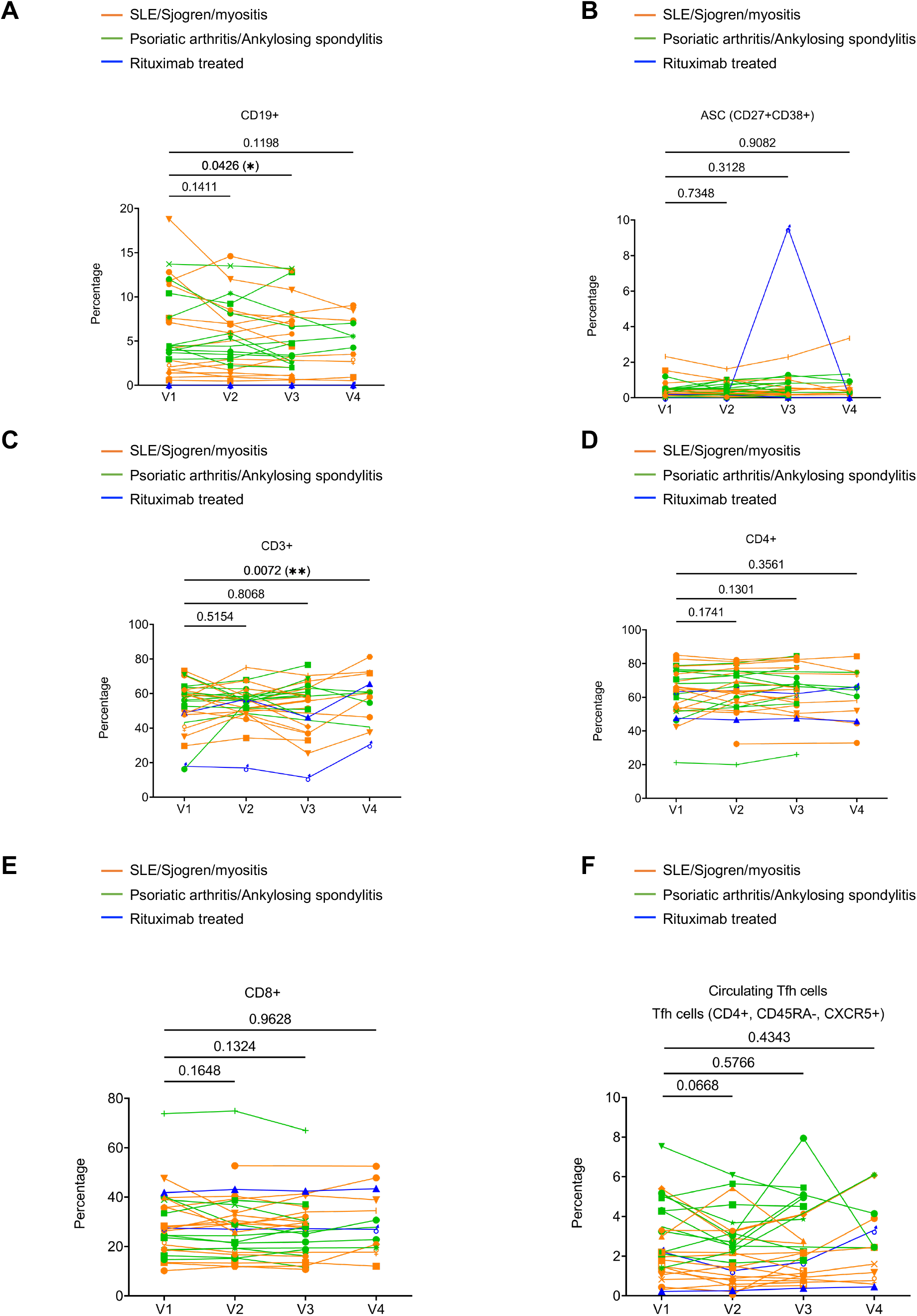
Summaries of the immune cell subset frequencies in rheumatic disease patients’ PBMC samples following SARS-CoV-2 mRNA vaccination. **A**, B cells (CD19^+^). **B**, Antibody secreting cells (ASC, CD19^+^CD27^+^CD38^+^). **C**, T cells (CD3^+^). **D**, CD4+ T cells (CD3^+^CD4^+^). **E**, CD8+ T cells (CD3^+^CD8^+^). **F**, Circulating follicular helper T cell (Tfh)-like cells (CD3^+^CD4^+^CD45RA^−^CXCR5^+^). *P* values were calculated by paired parametric t-test. Orange subgroup (n = 14). Green subgroup (n = 12). Blue subgroup (n = 2).

