## Supplementary Table 1-4 for "Immune responses and disease biomarker long-term changes following COVID-19 mRNA vaccination in a cohort of rheumatic disease patients"

**Supplementary Table 1.** Patients’ Demographics and Clinical Information.

|  | **Number of Patients** | **Percentage (%)** |
| --- | --- | --- |
| **Total** | **28** | **100** |
| **Gender** |  |  |
| Female | 20 | 71.42 |
| Male | 8 | 28.57 |
| **Age (years)** |  |  |
| Mean +/- sd | 53 (+/-12) |  |
| 20-29 | 2 | 7.14 |
| 30-39 | 2 | 7.14 |
| 40-49 | 7 | 25 |
| 50-59 | 8 | 28.57 |
| 60-69 | 7 | 25 |
| 70-79 | 1 | 3.57 |
| 80-89 | 1 | 3.57 |
| **Disease composition** |  |  |
| Systemic lupus erythematosus | 12 | 42.85 |
| Psoriatic Arthritis | 10 | 35.71 |
| Sjogren's syndrome | 4 | 14.28 |
| Idiopathic Inflammatory Myopathy | 1 | 3.57 |
| Ankylosing Spondylitis | 2 | 7.14 |
| **Visit** |  |  |
| 3 Visit | 15 | 53.57 |
| 4 Visit | 13 | 46.42 |
| **Vaccine** |  |  |
| PFIZER | 14 | 50 |
| MODERNA | 14 | 50 |

Note: One patient was diagnosed with both SLE and PsA.

**Supplementary Table 2.** Patients’ Medication exposure.

|  | **Number of Patients** | **Percentage (%)** | |
| --- | --- | --- | --- |
| **Total** | 28 | | 100 |
| **Prednisone** | 10 | | 35.71 |
| **Disease Modifying Antirheumatic Drug (DMARD)** |  | |  |
| Methotrexate | 6 | | 21.42 |
| Hydroxychloroquine | 11 | | 39.28 |
| Mycophenolate | 3 | | 10.71 |
| Azathioprine | 5 | | 17.85 |
| Leflunomide | 3 | | 10.71 |
| **Janus kinase inhibitors** |  | |  |
| Tofacitinib | 1 | | 3.57 |
| **Biologic therapies** |  | |  |
| Adalimumab | 2 | | 7.14 |
| Etanercept | 1 | | 3.57 |
| Rituximab | 2 | | 7.14 |
| Secukinumab | 4 | | 14.28 |
| Belimumab | 1 | | 3.57 |
| Ixekizumab | 1 | | 3.57 |
| **No DMARDs or biologics** | 3 | | 10.71 |

**Supplementary Table 3.** Primer sequences used for PCR.

| Name | Primers for PCR (5’-3’) |
| --- | --- |
| *MX1* | Forward GTTTCCGAAGTGGACATCGCA |
|  | Reverse CTGCACAGGTTGTTCTCAGC |
| *IFIT1* | Forward AGAAGCAGGCAATCACAGAAAA |
|  | Reverse CTGAAACCGACCATAGTGGAAAT |
| *IFI44* | Forward TGGTACATGTGGCTTTGCTC |
|  | Reverse CCACCGAGATGTCAGAAAGAG |

**Supplementary Table 4:** Antibodies used in Flow Cytometry Analysis

| **Antibody target** | **Fluorochrome conjugate** | **Clone** | **Supplier** | | **Catalog #** |
| --- | --- | --- | --- | --- | --- |
| CD56 | FITC | NCAM16.2 | BD Biosciences | 340723 | |
| CD20 | PE | 2H7 | Biolegend | | 302306 |
| HLA-DR | percp-cy5.5 | AC122 | Miltenyi Biotec | | 130095291 |
| viability | FVS 620 |  | BD Biosciences | | 564996 |
| CXCR5 | Alexa Fluor® 647 | J252D4 | Biolegend | | 356905 |
| CD4 | AF700 | RPA-T4 | Biolegend | | 300526 |
| CD45RA | APC-Cy7 | HI100 | Tonbo | | 25-0458-T100 |
| CCR7 | PE-Cy7 | G043H7 | Biolegend | | 353226 |
| CD38 | Pac-blue | HIT2 | Biolegend | | 303525 |
| CD8 | BV510 | RPA-T8 | Biolegend | | 301048 |
| CD127 | BV605 | A019D5 | Biolegend | | 351334 |
| CD3 | BV650 (Biolegend) | OKT3 | Biolegend | | 317324 |
| PD1 | BV711 | EH12.2H7 | Biolegend | | 329928 |
| CD25 | BV785 | BC96 | Biolegend | | 302638 |
| IgD | FITC | IA6-2 | Biolegend | | 348206 |
| CD24 | PE | ML5 | Biolegend | | 311106 |
| IgM | percp-cy5.5 | MHM-88 | Biolegend | | 314512 |
| CD21 | APC | B-ly4 | BD Biosciences | | 561357 |
| CD11c | RedFluor 710 | 3.9 | Tonbo Biosciences | | 80-0116 |
| CD138 | APC-Cy7 | MI15 | Biolegend | | 356528 |
| CD27 | PE-Cy7 | M-T271 | Biolegend | | 356412 |
| CD38 | BV421 | HIT2 | Biolegend | | 303525 |
| viability | BV510 |  | Tonbo Biosciences | | 13-0870-T100 |
| CD3 | SB600 | OKT3 | Invitrogen | | 63-0037-42 |
| CD19 | BV650 | HIB19 | Biolegend | | 302238 |
| CD14 | BV785 | M5E2 | Biolegend | | 301840 |
